# The iADJUST Study Protocol: A Randomised Waitlist-Controlled Feasibility Trial of iADJUST, a Digital Psychological Intervention for People Living with Chronic Kidney Disease

**DOI:** 10.64898/2026.06.29.26356845

**Authors:** Pooja Schmill, Joanna L Hudson, Sharlene Greenwood, Joseph Chilcot

## Abstract

**Background:** Psychological distress is common among people living with chronic kidney disease (CKD), yet access to structured psychological support within routine kidney care remains limited and varied. iADJUST is a primarily self-guided digital psychological intervention designed to support adjustment and emotional well-being in people living with CKD. Delivered over 12 weeks, iADJUST is hosted on the Kidney BEAM platform and supported by brief therapist contact to facilitate engagement. Before a definitive evaluation can be undertaken, it is necessary to establish the feasibility and acceptability of intervention delivery and study procedures.

**Aim:** To assess the feasibility and acceptability of delivering iADJUST to people living with CKD.

**Methods:** This study is a parallel-group, randomised, waitlist-controlled feasibility trial. Approximately 40-50 adults with CKD who are not receiving renal replacement therapy and have not received a kidney transplant will be randomised 1:1 to receive either iADJUST plus usual care or usual care alone. iADJUST is a six-session digital psychological intervention delivered over 12 weeks and supported by brief therapist contact. Feasibility outcomes include recruitment, retention, intervention uptake, engagement, completion of outcome measures, and acceptability of study procedures. Exploratory outcome measures include psychological distress, fatigue, functional impairment and quality of life collected to estimate variability, assess data completeness, and inform outcome selection for a future definitive trial. Following the primary endpoint assessment at 12 weeks, participants in both groups will receive access to the exercise-based Kidney BEAM programme. Kidney BEAM engagement and completion will be assessed at 24 weeks. The study is not powered to detect statistically significant between-group differences in clinical outcomes.

**Conclusion:** This study will provide evidence regarding the feasibility and acceptability of delivering iADJUST within UK kidney care pathways and inform the design of a future definitive trial.

**Trial Registration:** ISRCTN91507822

## Introduction

Psychological well-being is increasingly recognised as an important component of comprehensive kidney care [1, 2]. Despite this, access to structured psychological support remains inconsistent across kidney services within the UK National Health Service (NHS) [3], particularly for individuals earlier in the disease pathway. Consequently, many people experiencing emotional difficulties associated with living with CKD receive limited structured psychological support beyond routine clinical care [2].

Digital interventions have emerged as a potential approach to increasing access to support while reducing barriers associated with healthcare access, fluctuating symptoms, fatigue, and service capacity constraints [4, 5]. Digitally delivered programmes are increasingly being incorporated into long-term condition management pathways to support self-management and improve access to care [5, 6].

Kidney BEAM is a physical-activity-based digital health intervention developed specifically for people living with kidney disease. Commissioned by several NHS Trusts in the UK, Kidney BEAM provides access to online physical activity, self-management, and well-being resources and has demonstrated high levels of engagement and acceptability among people living with CKD [7]. Emerging evidence suggests that participation in Kidney BEAM may support physical activity, health-related quality of life, and self-management behaviours, highlighting the potential value and cost-effectiveness of digitally delivered interventions within kidney care pathways [7, 8]. Kidney BEAM, therefore, provides an appropriate platform through which to deliver and evaluate novel CKD-specific psychosocial interventions such as iADJUST.

iADJUST is a primarily self-guided digital psychological intervention designed to support adjustment and emotional well-being in people living with CKD and hosted on the Kidney BEAM digital platform. The intervention was developed through a systematic evidence-, theory-, and patient-informed process, which is described in detail elsewhere [9]. iADJUST aims to support psychological adjustment to living with CKD through a series of self-directed digital sessions delivered over 12 weeks and supported by brief therapist contact to promote engagement and intervention competition.

Before conducting a definitive trial, it is necessary to establish the feasibility and acceptability of both the intervention and trial procedures, including recruitment, retention, intervention engagement, and outcome data collection. Key uncertainties include recruitment and retention rates, intervention engagement and acceptability of study procedures, and completion of outcome measures. The present study has therefore been designed as a randomised feasibility trial to evaluate these parameters and inform the design of a future definitive trial.

### Aim

To assess the feasibility and acceptability of delivering iADJUST, a digital psychological intervention designed to support adjustment and emotional well-being in CKD.

### Objectives

#### Feasibility Objectives

- To assess the feasibility of recruitment to a randomised feasibility trial of iADJUST.
- To assess participant retention throughout the study.
- To assess the acceptability of study procedures, including consent, randomisation, and outcome data collection.
- To assess intervention uptake, engagement and completion.
- To assess the feasibility of delivering iADJUST remotely through the Kidney BEAM platform
- To estimate the completeness and variability of selected patient-reported outcome measures.
- To describe uptake and engagement with the Kidney BEAM programme after completing iADJUST.
- To inform progression decisions and the design of a future definitive trial.

## Methods

### Study Design

This study is a parallel-group, randomised, waitlist-controlled feasibility trial. Following informed consent and completion of baseline assessments, participants will be randomised in a 1:1 ratio using a secure computer-generated allocation sequence to either:

- iADJUST plus usual care
- Usual care (waitlist control)

Participants allocated to iADJUST will receive immediate access to the intervention for 12 weeks in addition to usual kidney care. Participants allocated to the wait-listed control arm will continue to receive usual kidney care during this period.

Following completion of the 12-week assessment, participants in both groups will receive access to the Kidney BEAM programme. Following completion of the final 24-week assessment, participants allocated to the control arm will be offered access to iADJUST.

The study will be conducted and reported in accordance with CONSORT extension guidance for pilot and feasibility trials [10] The protocol has been developed in accordance with the SPIRIT reporting guidelines [11].

### Study Setting

Participants will be recruited from NHS kidney services in the UK that are involved in the Kidney BEAM trial [7].

### Patient and Public Involvement and Engagement (PPIE)

People living with CKD contributed to the development and refinement of iADJUST through a structured PPIE process. Contributors provided feedback on intervention content, language, delivery format, session structure, and acceptability. Their feedback informed the refinement of intervention materials and study procedures. Details of the intervention development process are described elsewhere [9]. Contributors are not involved in participant recruitment or data analysis for the present feasibility study.

### Ethics and Registration

iADJUST is a sub-study of the Kidney BEAM trial (NCT04872933) and is sponsored by King’s College Hospital NHS Foundation Trust. The study has received favourable ethical approval from the Bromley Research Ethics Committee (REC Reference: 21/LO/0243) and approval from the Health Research Authority (IRAS No: 291403). This trial was prospectively registered with the [ISRCTN Registry](https://www.isrctn.com/) (Registration No: ISRCTN91507822). Any protocol amendments will be submitted to the sponsor, Research Ethics Committee and Health Research Authority for approval prior to implementation where required. Relevant amendments will also be updated in the ISRCTN trial registry.

### Trial Governance

Pooja Schmill is the Chief Investigator and is responsible for day-to-day trial management, participant recruitment, intervention delivery, data collection, and analysis. Professor Joseph Chilcot, Professor Sharlene Greenwood and Dr Joanna Hudson provide scientific and clinical oversight throughout the study. King’s College Hospital NHS Foundation Trust is the study sponsor.

### Sample Size

The study aims to recruit approximately 40–50 participants. Consistent with recommendations for feasibility studies, the sample size has not been determined based on statistical power but is intended to provide preliminary estimates of recruitment, retention, intervention engagement, and outcome completion to inform a future definitive trial [12, 13].

### Eligibility Criteria

Trial eligibility criteria are summarised in Table 1.

**Table 1.** Inclusion and exclusion criteria.

| Inclusion Criteria | Exclusion Criteria |
| --- | --- |
| Adults aged $\geq 18$ years | Acute kidney injury without established CKD |
| Diagnosis of CKD | Current renal replacement therapy or previous kidney transplant |
| Access to an internet-enabled device | Receipt of structured psychological therapy within the previous three months |
| Able to engage with a self-guided, digital programme | Participation in Kidney BEAM or structured exercise programme within the previous three months |
| Able to understand written & spoken English | Current severe mental illness or psychiatric instability judged to make participation unsuitable, including recent suicidal ideation, suicide attempt, or active risk requiring urgent mental health intervention |
| Able to provide informed consent | Severe cognitive, sensory (e.g., visual or hearing), communication, or other impairments, or medical conditions judged by the clinical team or Principal Investigator to make participation inappropriate |

### Consent Procedures

Eligible participants will receive a Participant Information Sheet and have the opportunity to discuss the study with a member of the research team. Participants will provide electronic informed consent before enrolment and randomisation.

### Randomisation & Allocation Concealment

Following completion of baseline assessments, authorised members of the research team will enrol participants and obtain allocation through the Sealed Envelope randomisation service which is a secure web-based randomisation service (e.g., Sealed Envelope) employing a computer-generated allocation sequence. Allocation concealment will be maintained through the Sealed Envelope system, with treatment allocation revealed only after participant enrolment and completion of baseline assessments.

### Blinding

Due to the nature of the intervention, participants and members of the research team delivering brief therapist contacts cannot be blinded to treatment allocation. Outcome measures are self-reported and collected electronically; consequently, outcome assessors and participants cannot be blinded.

### Intervention

iADJUST is a six-session digital psychological intervention delivered through the Kidney BEAM platform over approximately 12 weeks. The programme includes psychoeducation, guided audio reflections, behavioural strategies, and self-directed activities designed to support psychological adjustment and emotional well-being in people living with CKD. The six sessions are:

1. Understanding Life with CKD
2. Managing Fatigue and Energy
3. Managing Worry and Living with Uncertainty
4. Understanding the Inner Critical Voice
5. Managing Mood and Staying Active
6. Maintaining Progress and Looking Ahead

Participants allocated to iADJUST will receive access to the intervention for 12 weeks alongside brief therapist contact delivered remotely by trained members of the research team, including trainee psychologists. Support contacts are intended to facilitate engagement with programme content, support goal setting, encourage ongoing participation and intervention completion, and help address practical barriers to intervention use. These contacts are not intended to provide additional psychological treatment beyond the intervention itself.

Participants may withdraw from the intervention or study at any time without affecting their usual NHS care. The research team may discontinue participation if continued involvement is considered inappropriate due to safeguarding concerns or withdrawal of consent.

### Comparator

Participants allocated to the control arm will continue to receive usual kidney care during the first 12 weeks of the study. A waitlist-control design was selected to assess feasibility while ensuring that all participants had the opportunity to access the intervention content.

Participants in both study arms will continue to receive routine NHS kidney care throughout the study. Access to other healthcare services will not be restricted.

### Outcome Measures

#### Primary Feasibility Outcomes

The primary feasibility outcomes will be assessed at 12 weeks (the primary endpoint) and will include:

- Recruitment rate
- Consent rate
- Retention at 12-weeks
- Completion of outcome measures
- iADJUST uptake
- iADJUST engagement
- Kidney BEAM uptake

#### Definitions

- **iADJUST uptake:** Commencement of at least one iADJUST session.
- **iADJUST engagement:** Completion of a minimum of four of the six iADJUST sessions.
- **Full intervention completion:** Completion of all six iADJUST sessions.
- **Kidney BEAM uptake:** Registration for the Kidney BEAM programme following completion of the 12-week intervention period.

#### Secondary Feasibility Outcomes (24-week follow-up)

The following secondary feasibility outcomes will be assessed at 24 weeks:

1. Retention at final follow-up.
2. Kidney BEAM engagement metrics were obtained from routinely collected platform usage data.

#### Exploratory Outcomes

The following patient-reported outcome measures will be collected at baseline and the primary endpoint (12-week) to assess data completeness and estimate variability for a future definitive trial.

1. **Depression symptoms** will be assessed using the Patient Health Questionnaire-8 (PHQ-8), an eight-item measure of depressive symptoms with scores ranging from 0 to 24 [14]. The PHQ-8 has demonstrated comparable psychometric properties to the PHQ-9 in population and clinical samples [15].
2. **Anxiety symptoms** will be assessed using the Generalised Anxiety Disorder-7 (GAD-7), a seven-item measure of anxiety symptoms with scores ranging from 0–21 [16].
3. **Psychological distress** will be assessed using a PHQ-ADS composite score (0-45) derived from PHQ-8 and GAD-7 responses. The PHQ-ADS captures the combined burden of depressive and anxiety symptoms and has been validated in people living with long-term conditions, where these symptoms frequently co-occur [14].
4. **Fatigue** will be assessed using the Chalder Fatigue Questionnaire (CFQ), a widely used measure of physical and mental fatigue. Scores range from 0–33 [17].
5. **Functional impairment** will be assessed using the Work and Social Adjustment Scale (WSAS; score range 0–40), a five-item measure of impairment in work, home management, social leisure, private leisure, and close relationships [18].
6. **Health-related quality of life** will be assessed using the 12-Item Short Form Health Survey (SF-12), a widely used measure of physical and mental health-related quality of life [19].

Higher scores indicate greater symptom severity or impairment across all measures except SF-12, where higher scores indicate better perceived health status.

#### Participant Timeline

The participant timeline and study flow are summarised in Fig 1.

**Fig 1.**
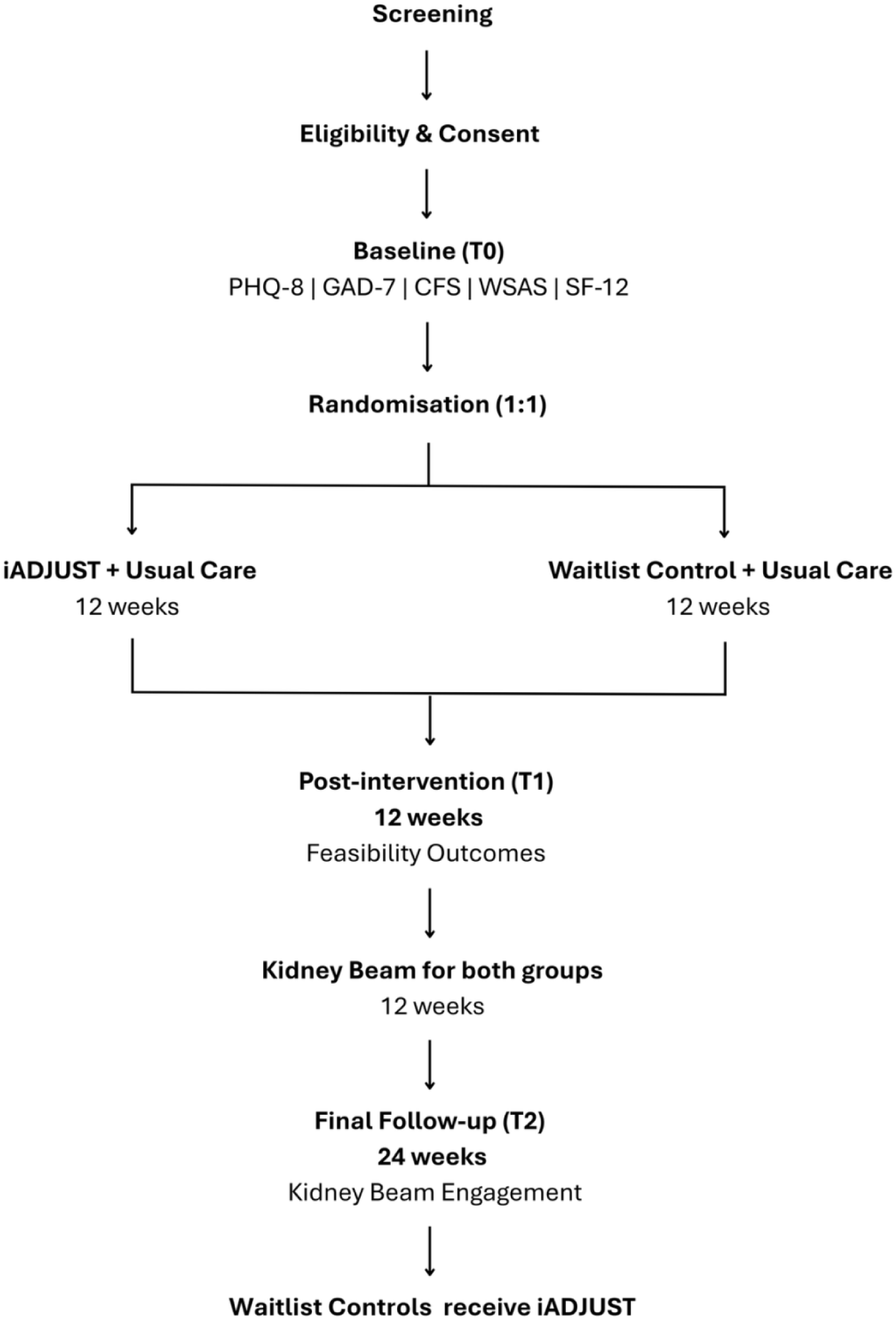
Study flow and participant timeline.

#### Schedule of assessments

The schedule of enrolment, interventions, and assessments is summarised in Table 2 in accordance with SPIRIT guidance. Participants will receive reminder emails to complete follow-up assessments. Where appropriate, additional reminders may be sent to maximise follow-up data completeness.

**Table 2.** Schedule of enrolment, interventions, and assessment.

| Procedure | Screening | Baseline (T0) | Week 12 (T1) | Week 24 (T2) |
| --- | --- | --- | --- | --- |
| Informed consent | X |  |  |  |
| Eligibility assessment | X |  |  |  |
| Randomisation |  | X |  |  |
| Demographic and clinical data |  | X | X |  |
| PHQ-8 |  | X | X |  |
| GAD-7 |  | X | X |  |
| Chalder Fatigue Questionnaire |  | X | X |  |
| WSAS |  | X | X |  |
| SF-12 |  | X | X |  |
| iADJUST engagement/completion metrics |  |  | X |  |
| Kidney BEAM registration/uptake |  |  | X |  |
| Kidney BEAM engagement metrics |  |  |  | X |

#### Progression Criteria

Progression criteria were specified a priori using a traffic-light framework to inform decisions regarding progression to a future definitive trial. The selected domains reflect commonly recommended indicators of feasibility, including recruitment, retention, intervention uptake, engagement, and outcome completion, which are among the most frequently reported progression criteria in pilot and feasibility trials [12, 20, 21].

Thresholds were selected pragmatically to reflect the anticipated challenges associated with delivering a novel digital psychological intervention to people living with CKD earlier in the kidney care pathway. Retention and outcome completion thresholds were set at ≥70% for progression, consistent with commonly used benchmarks in feasibility studies, empirical work examining progression criteria thresholds, and recommendations that follow-up rates below 50% may substantially limit interpretation of feasibility outcomes [20, 22, 23]. Intervention uptake and adherence thresholds were informed by evidence demonstrating substantial variability in engagement with digital psychological interventions and the recognition that intervention engagement is not solely reflected by programme completion [24, 25]. Full intervention completion thresholds were therefore intentionally more conservative than uptake and engagement.

Feasibility will be assessed using the traffic-light framework presented in Table 3. Progression decisions will be based on an overall assessment of feasibility findings rather than any single criterion and will consider recruitment, retention, intervention engagement, outcome completion, and acceptability of study procedures collectively.

**Table 3.** Feasibility Assessment C.

| Domain | Green (Proceed) | Amber (Amend) | Red (Reconsider) |
| --- | --- | --- | --- |
| Recruitment | $\geq 60\%$ | 40–59% | $< 40\%$ |
| Retention | $\geq 70\%$ | 50–69% | $< 50\%$ |
| Outcome completion | $\geq 70\%$ | 50–69% | $< 50\%$ |
| Uptake | $\geq 60\%$ | 40–59% | $< 40\%$ |
| Engagement ($\geq 4$ sessions) | $\geq 50\%$ | 30–49% | $< 30\%$ |
| Full intervention completion* | $\geq 30\%$ | 15–29% | $< 15\%$ |
| *\*Full intervention completion refers to completion of all six iADJUST sessions.* |  |  |  |

### Data Analysis

Analyses will follow the intention-to-treat principle, with participants analysed according to their allocated study group where outcome data are available. Analyses will be primarily descriptive and will focus on estimation rather than hypothesis testing, consistent with recommendations for feasibility studies. Continuous variables will be summarised using means and standard deviations or medians and interquartile ranges as appropriate. Categorical variables will be summarised using frequencies and percentages.

The study is not powered to detect statistically significant between-group differences in clinical outcomes. Missing data will not be imputed. Rates and patterns of missing data will be summarised descriptively to inform the design of a future definitive trial. For the exploratory continuous outcomes at week 12 (T1), effect sizes with 95% confidence intervals will be estimated using linear regression models, with group allocation and the baseline value of the outcome included as covariates.

### Data Management

Study procedures will comply with the UK General Data Protection Regulation (GDPR), the Data Protection Act 2018, and sponsor governance requirements. Participant data will be pseudonymised and stored securely on password-protected systems accessible only to authorised members of the research team. Participant confidentiality will be maintained throughout the study and identifiable information will be accessible only to authorised members of the research team.

### Harms / Adverse Events

Any adverse events, participant concerns, or safeguarding issues identified during the study will be documented and managed in accordance with sponsor and NHS governance procedures. Participants experiencing significant distress will be signposted to appropriate healthcare services, including their GP or renal care team, where appropriate.

### Trial Monitoring

Given the low-risk nature of the study, a formal Data Monitoring Committee was not considered necessary. Trial conduct will be overseen by the study investigators and sponsor in accordance with institutional governance procedures.

### Data Sharing

De-identified participant data may be made available from the corresponding author upon reasonable request following publication of the main study findings, subject to institutional and ethical approvals.

## Discussion

This study will provide important evidence regarding the feasibility and acceptability of delivering iADJUST within kidney care pathways. Findings will inform progression to a future definitive trial and contribute to the evidence base for digital psychological interventions delivered earlier in the CKD pathway. The findings will also help determine whether a future definitive trial of iADJUST is feasible and acceptable within routine kidney care pathways.

## Dissemination

Study findings will be disseminated through peer-reviewed publications, conference presentations, and other academic dissemination activities. Participants may request a summary of study findings following study completion.

## Data Availability

All data produced in the present work are contained in the manuscript

## Acknowledgements

The authors would like to thank the people living with chronic kidney disease who contributed to the development and refinement of iADJUST through patient and public involvement activities. Their insights and feedback helped inform the intervention content, delivery, and study procedures.

## Protocol Contributors

Pooja Schmill conceived the study, developed the intervention, designed the trial protocol, and is responsible for the day-to-day conduct of the study. Professor Joseph Chilcot, Professor Sharlene Greenwood, and Dr Joanna Hudson contributed to study conception, protocol development, intervention design, and provided scientific and clinical supervision throughout the study. All authors reviewed, revised, and approved the final protocol.

## References

1. Chilcot, J., et al., Mental health and kidney disease. Nature Reviews Nephrology, 2026. 22(5): p. 363–376.

2. UK, K.R., Addressing the mental health challenges of life with kidney disease: The case for change (J. Wilton). 2023.

3. Chilcot, J., et al., The identification and management of depression in UK Kidney Care: Results from the Mood Maps Study. J Ren Care, 2024. 50(3): p. 297–306.

4. Al Mahmud, A., et al., Digital Health Interventions to Support Chronic Disease Management: Systematic Scoping Review. JMIR Mhealth Uhealth, 2026. 14: p. e63742.

5. Zheng, X., et al., Digital symptom management interventions for people with chronic kidney disease: a scoping review based on the UK Medical Research Council Framework. BMC Public Health, 2024. 24(1): p. 3534.

6. Lightfoot, C.J., et al., The effects of a digital health intervention on patient activation in chronic kidney disease. npj Digital Medicine, 2024. 7(1): p. 318.

7. Greenwood, S.A., et al., Evaluating the effect of a digital health intervention to enhance physical activity in people with chronic kidney disease (Kidney BEAM): a multicentre, randomised controlled trial in the UK. The Lancet Digital Health, 2024. 6(1): p. e23–e32.

8. Greenwood, S.A., et al., Kidney Beam-A Cost-Effective Digital Intervention to Improve Mental Health. Kidney International Reports, 2024. 9(11): p. 3204–3217.

9. Schmill, P., et al., Development of iADJUST: a theory-informed, patient co-designed digital psychological intervention for adjustment in chronic kidney disease. medRxiv, 2026: p. 2026.06.10.26355356.

10. Hopewell, S., et al., CONSORT 2025 statement: updated guideline for reporting randomised trials. Lancet, 2025.

11. Hróbjartsson, A., et al., SPIRIT 2025 explanation and elaboration: updated guideline for protocols of randomised trials. Bmj, 2025. 389: p. e081660.

12. Eldridge, S.M., et al., Defining Feasibility and Pilot Studies in Preparation for Randomised Controlled Trials: Development of a Conceptual Framework. PLoS One, 2016. 11(3): p. e0150205.

13. Sim, J. and M. Lewis, The size of a pilot study for a clinical trial should be calculated in relation to considerations of precision and efficiency. J Clin Epidemiol, 2012. 65(3): p. 301–8.

14. Kroenke, K., et al., Patient Health Questionnaire Anxiety and Depression Scale: Initial Validation in Three Clinical Trials. Psychosom Med, 2016. 78(6): p. 716–27.

15. Wu, Y., et al., Equivalency of the diagnostic accuracy of the PHQ-8 and PHQ-9: a systematic review and individual participant data meta-analysis. Psychological Medicine, 2020. 50(8): p. 1368–1380.

16. Spitzer, R.L., et al., A brief measure for assessing generalized anxiety disorder: the GAD-7. Arch Intern Med, 2006. 166(10): p. 1092–7.

17. Chalder, T., et al., Development of a fatigue scale. J Psychosom Res, 1993. 37(2): p. 147–53.

18. Mundt, J.C., et al., The Work and Social Adjustment Scale: a simple measure of impairment in functioning. Br J Psychiatry, 2002. 180: p. 461–4.

19. Ware, J., Jr., M. Kosinski, and S.D. Keller, A 12-Item Short-Form Health Survey: construction of scales and preliminary tests of reliability and validity. Med Care, 1996. 34(3): p. 220–33.

20. Avery, K.N.L., et al., Informing efficient randomised controlled trials: exploration of challenges in developing progression criteria for internal pilot studies. BMJ Open, 2017. 7(2): p. e013537.

21. Mellor, K., et al., Recommendations for progression criteria during external randomised pilot trial design, conduct, analysis and reporting. Pilot and Feasibility Studies, 2023. 9(1): p. 59.

22. Bell, M.L., A.L. Whitehead, and S.A. Julious, Guidance for using pilot studies to inform the design of intervention trials with continuous outcomes. Clin Epidemiol, 2018. 10: p. 153–157.

23. Mbuagbaw, L., et al., Empirical progression criteria thresholds for feasibility outcomes in HIV clinical trials: a methodological study. Pilot Feasibility Stud, 2023. 9(1): p. 96.

24. Donkin, L., et al., A systematic review of the impact of adherence on the effectiveness of e-therapies. J Med Internet Res, 2011. 13(3): p. e52.

25. Perski, O., et al., Conceptualising engagement with digital behaviour change interventions: a systematic review using principles from critical interpretive synthesis. Transl Behav Med, 2017. 7(2): p. 254–267.

